# Health Insurance Expenditure Structure for Type 2 Diabetes Treatment in Vietnam by Hospital Classification, 2018–2022: A Descriptive Analysis of Claims Data From Hanoi and Ho Chi Minh City

**DOI:** 10.64898/2026.04.09.26350559

**Authors:** Thuy Thi Thu Nguyen, Viet Linh Nguyen, Nhi Ngoc Yen Vo, Huy Cao Duc Nguyen, Huong Thi Thanh Nguyen

**Author notes:** Correspondence: Author: Thuy Thi Thu Nguyen.

## Abstract

**Background:** Type 2 diabetes mellitus (T2DM) is a chronic disease that imposes a significant burden on healthcare systems and society. In Vietnam, the prevalence of T2DM is rapidly increasing; however, evidence on treatment expenditure derived from large administrative databases remains limited. This study was carried out provides an overview of total treatment expenditures for T2DM across hospital tiers between 2018 and 2022.

**Methods:** This cross-sectional descriptive study utilized retrospective health insurance (HI) data from 2018–2022. Data was collected and analyzed based on cost components (medications, diagnostic tests, hospital beds, etc.) across healthcare facilities classified by hospital level. Costs were converted to 2024 USD using the CCEMG–EPPI-Centre cost converter.

**Results:** Total expenditure increased from 227.17 million USD in 2018 to 425.53 million USD in 2022 with spending concentrated in Class I and Class II healthcare facilities, although their shares declined over time, while the proportions attributed to unclassified and special-class facilities increased. Drugs accounted for the largest share of expenditure (49.65%–78.95%), followed by laboratory tests (7.31%–19.89%) across all hospital classifications. Other components, including hospital beds, diagnostic imaging, procedures/surgeries, and medical supplies, contributed smaller proportions but increased over time in several facility groups.

**Conclusion:** The study indicates that medication costs constitute the largest share of treatment expenditure for type 2 diabetes mellitus at healthcare facilities, reflecting the long-term treatment requirements of this chronic disease. In addition, health expenditure remained concentrated in Class I and Class II healthcare facilities, although their shares declined over the study period, while the proportions attributed to unclassified and special-class facilities increased. These findings suggest the need to strengthen diabetes screening, treatment, and follow-up at lower-level healthcare facilities in order to reduce the burden on higher-level hospitals and improve the efficiency of healthcare resource allocation.

## 1. Introduction

Diabetes Mellitus (DM) is one of the most common non-communicable diseases in both developing and developed countries. Numerous studies have indicated a high prevalence of diabetes in countries with low and middle-income economies (1). DM imposes a significant burden on society, including increased direct medical costs, loss of labor productivity, premature mortality, and intangible costs such as reduced social connectivity and quality of life (2). According to the International Diabetes Federation (IDF), in 2021, there were 537 million people (aged 20–79) worldwide living with DM, and this number is expected to rise to 643 million by 2030 and 783 million by 2045. Additionally, the IDF estimates that over 6.7 million people in the 20–79 age group died from diabetes-related causes in 2021 (3).

The hospital system in Vietnam is classified to standardize healthcare service quality, optimize management, and allocate medical resources effectively. This classification is regulated by the Ministry of Health based on criteria such as bed capacity, professional qualifications, infrastructure, scope of medical services, and quality of patient care. Hospitals in Vietnam are categorized into special class, Class I, Class II, Class III, Class IV, and unclassified hospitals. Special-class hospitals are the largest medical institutions which specialize in medical services, conduct scientific research, train healthcare personnel with over 1,000 inpatient beds, and offer technical support to lower-level healthcare facilities. Class I hospitals have a capacity ranging from 500 to 1,000 beds and are equipped to perform advanced medical procedures, specialized care, and engage in research and professional training. Class II hospitals are medium-sized facilities with 300 to 500 beds, they provide common healthcare services, emergency care, inpatient treatment, and perform intermediate-level medical procedures under the guidance of higher-level hospitals. Class III hospitals have a bed capacity of 100 to 300, mainly serving as district-level hospitals or medical centers. These hospitals focus on primary healthcare, emergency stabilization, and patient referrals to higher-level facilities when necessary. Class IV hospitals are small-scale facilities with fewer than 100 beds, they are the most basic level of hospital facilities which offer essential healthcare services, basic medical treatment, and community health support. Additionally, there are unclassified hospitals, which include facilities that do not meet the official classification standards or small private hospitals. These institutions typically provide limited medical services and do not engage in advanced research or professional training.

In Vietnam, according to the IDF in 2021, it is estimated that nearly 4 million people are living with DM, and this number is expected to continue rising, reaching approximately 6 million by 2045 (4). Although several studies have described diabetes-related costs, Vietnamese evidence is still largely based on data from single hospitals or limited local samples. Accordingly, this study aimed to describe health insurance expenditure for T2DM treatment in Hanoi and Ho Chi Minh City using Social Health Insurance in payment data from 2018 to 2022, with results presented by hospital classification and expenditure component.

## 2. Methods

### 2.1. Study setting and design

A descriptive cross-sectional study was conducted using retrospective electronic payment data from the Social Health Insurance (SHI) systems of Ho Chi Minh City and Hanoi for the period 2018–2022. Secondary data were obtained from the Social Security agencies in Hanoi and Ho Chi Minh City. The Hanoi dataset was accessed for research purposes on January 20, 2023, and the Ho Chi Minh City dataset was accessed on March 16, 2023. During data acquisition, the authors had access to potentially identifiable participant information. All data were managed under strict confidentiality requirements and were used exclusively for research purposes.

### 2.2. Study sample

All records that met the inclusion criteria and did not meet any of the exclusion criteria during the sampling period were included.

Inclusion criteria and exclusion criteria are presented in **Table 1**.

**Table 1.** Inclusion criteria and exclusion criteria.

| Inclusion criteria | Exclusion criteria |
| --- | --- |
| - Type 2 diabetes patients (ICD 10: E11) applied in the primary diagnosis field.<br>- Patients enrolled in HI and utilizing it for the treatment | - Patients lacking necessary data for the study. |

*Notes: ICD – International Statistical Classification of Diseases and Related Health Problems; HI - Health Insurance.*

### 2.3. Study variables

The primary outcome was total direct medical expenditure for T2DM treatment in the eligible SHI population from 2018 to 2022. Expenditure was described according to absolute amount and relative share of each component. Study variables included:

- Demographic characteristics: Gender (male/female), age group (18-39 years/40-59 years/over 60 years), age.

- Disease characteristics: comorbidities (non HTN (hypertension) and DLD (dyslipidemia)/ HTN + DLD/DLD/HTN), type of complications (LBV: Large Blood Vessels/SBV: Small Blood Vessels/MC: Metabolic Complications/LBV + SBV: Large Blood Vessels + Small Blood Vessels/LBV + MC: Large Blood Vessels + Metabolic Complications/SBV + MC: Small Blood Vessels + Metabolic Complications/LBV + SBV + MC: Large Blood Vessels + Small Blood Vessels + Metabolic Complications), average number of inpatient treatment days per treatment episode.

- Expenditure components: drug expenditure, laboratory tests expenditure, medical consultation expenditure, medical bed expenditure, medical imaging diagnosis expenditure, surgical procedures expenditure, medical supplies expenditure, blood expenditure, technical services expenditure, transportation expenditure.

### 2.4. Statistical Methods and Data Processing

The direct medical costs are converted to USD in 2024 by using the tool of CCEMG - EPPI-Centre Cost Converter.

Data collected in the study were processed using Microsoft Excel and the R statistical software, with statistical confidence set at 95%. Complications were identified based on the study by Young et al. (5) with conversion to ICD-10 codes according to the mapping developed by Glasheen et al. (6). Comorbidities were determined based on ICD-10 codes, including dyslipidemia (E78) and hypertension (I10–I15). The analysis was descriptive only: categorical variables were summarized as counts and percentages, while expenditure totals and component shares were summarized by year and hospital classification.

## 3. Results

### 3.1. Characteristics of the study sample

The characteristics of the study sample, classified by the classification of healthcare facilities (HCF) from 2018 to 2022, are presented in **Table 2**.

[Table 2 was uploaded separately and should appear here].

In terms of demographic characteristics, the study recorded an average annual male-to-female ratio of approximately 1:1.24 across all HCFs from 2018 to 2022, indicating a higher proportion of females than males. This pattern was observed across all HCF classifications except for special-class HCFs, where the ratio was reversed (male-to-female ratio of 1:0.69). Additionally, the study found that the average annual number of patients aged 60 years and above was the highest across all HCFs (351,921 patients), followed by patients aged 40–59 years (193,735 patients), while the number of patients aged 18–39 years was negligible (24,608 patients). The ranking of age subgroups was similarly observed within each HCF classification.

Regarding comorbidities, the annual average number of patients with both DLD and HTN accounted for the largest share among comorbidity groups across all HCF classifications (349,222 patients, representing 61.24%). This was observed in most HCF classifications, ranging from 28.47% (Fourth-class HCFs) to 65.48% (First-class HCFs), except in Class IV HCFs, where the annual average number of patients with HTN alone represented the highest proportion (31.84%). Meanwhile, the annual average number of patients with HTN alone accounted for the lowest proportion among patients with comorbidities across all HCF classifications (51,390 patients, representing 9.01%), except for Class IV HCFs (ranked first) and special-class HCFs (ranked third).

Additionally, the annual average number of patients with LBV accounted for the majority of cases across all HCF classifications (210,665 patients, representing 36.94%). Within individual HCF classifications, this proportion ranged from 23.05% (Fourth-class HCFs) to 39.44% (First-class HCFs). Patients with both LBV and SBV represented the second-largest annual average group among all HCFs (106,430 patients, accounting for 18.66%). This trend was consistent across most HCF classifications, ranging from 7.65% (special-class HCFs) to 21.48% (unclassified HCFs), except for special-class HCFs, where patients with SVC ranked second (8.10%). Furthermore, the study recorded an overall average inpatient treatment duration of 45.82 days per episode across all HCF classifications.

### 3.2. Results of the analysis of the total expenditure for type 2 diabetes treatment

The results of the analysis of the total expenditure for T2DM treatment over the entire period from 2018 to 2022 are presented in **Table 3**.

**Table 3.** Total expenditure for type 2 diabetes treatment from 2018 to 2022 (million USD)

|  | **2018** | **2019** | **2020** | **2021** | **2022** |
| --- | --- | --- | --- | --- | --- |
| Drugs | 146.06 | 198.69 | 219.26 | 199.67 | 245.74 |
|  | (64.2968%) | (60.9115%) | (59.7752%) | (59.9842%) | (57.7501%) |
| Laboratory Tests | 35.05<br>(15.4296%) | 47.17<br>(14.1188%) | 47.85<br>(13.0448%) | 42.30<br>(12.7086%) | 54.10<br>(12.7124%) |
| Medical Consultation | 11.56<br>(5.0864%) | 15.28<br>(4.5720%) | 15.02<br>(4.0942%) | 11.68<br>(3.5103%) | 14.68<br>(3.4511%) |
| Hospital Bed | 16.62<br>(7.3161%) | 27.56<br>(8.2475%) | 27.82<br>(7.5836%) | 23.23<br>(6.9777%) | 31.04<br>(7.2943%) |
| Diagnostic imaging | 8.98<br>(3.9517%) | 14.02<br>(4.1954%) | 16.34<br>(4.4548%) | 15.39<br>(4.6226%) | 21.10<br>(4.9569%) |
| Procedures and surgeries | 5.47<br>(2.4079%) | 11.68<br>(3.4961%) | 13.97<br>(3.8084%) | 14.38<br>(4.3188%) | 22.41<br>(5.2664%) |
| Medical Supplies | 2.55<br>(1.1213%) | 12.91<br>(3.8640%) | 23.63<br>(6.4418%) | 22.99<br>(6.9056%) | 32.68<br>(7.6806%) |
| Blood products | 0.65<br>(0.1011%) | 1.43<br>(0.4275%) | 2.11<br>(0.5741%) | 2.20<br>(0.6619%) | 3.47<br>(0.8149%) |
| Technical Services | 0.29<br>(0.0021%) | 0.56<br>(0.1665%) | 0.82<br>(0.228%) | 1.03<br>(0.3097%) | 0.32<br>(0.0729%) |
| Transportation | 0.004<br>(0.0021%) | 0.003<br>(0.0008%) | 0.001<br>(0.0005%) | 0.002<br>(0.0007%) | 0.002<br>(0.0006%) |
| **Total expenditure** | **227.17** | **326.19** | **366.81** | **332.87** | **425.53** |

**Table 4.**
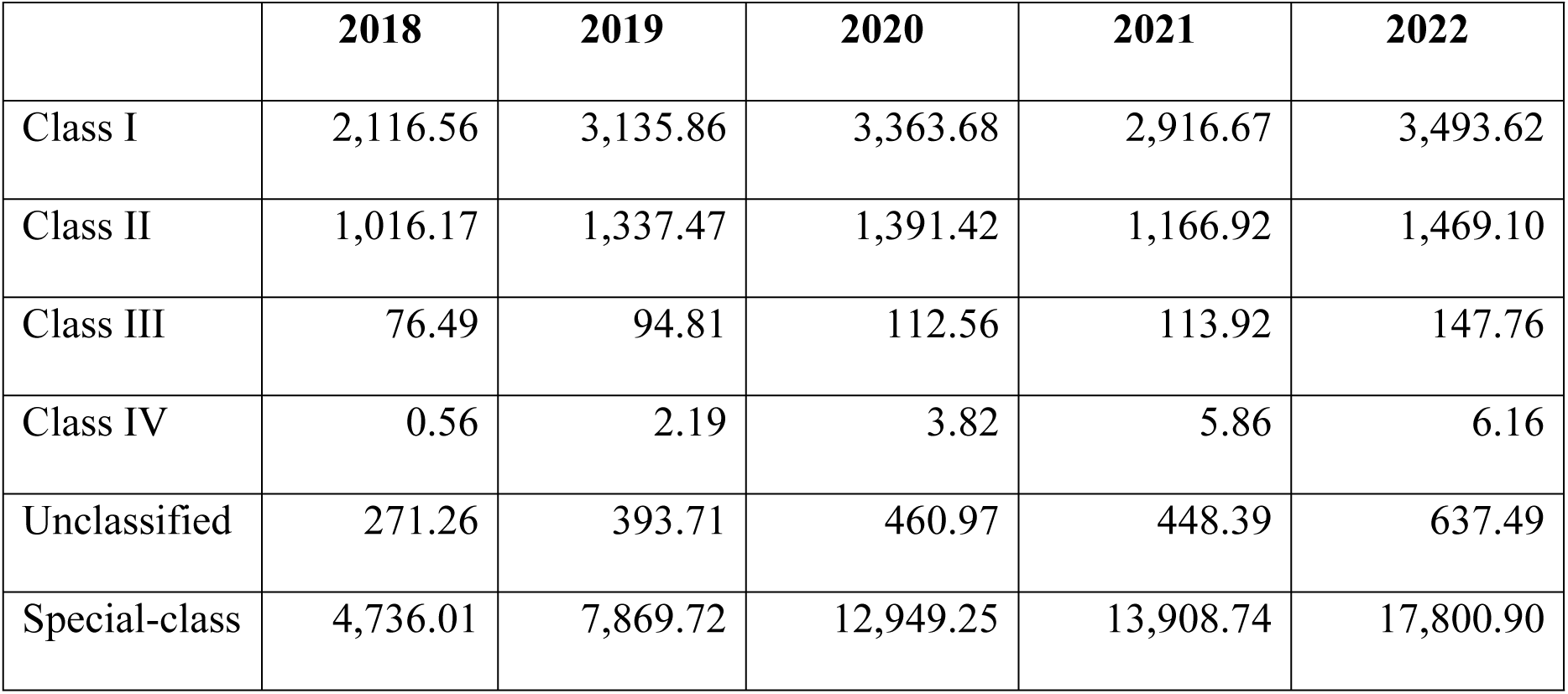
Average health expenditure for type 2 diabetes treatment at each hospital from 2018 to 2022 (thousand USD)

|  | **2018** | **2019** | **2020** | **2021** | **2022** |
| --- | --- | --- | --- | --- | --- |
| Class I | 2,116.56 | 3,135.86 | 3,363.68 | 2,916.67 | 3,493.62 |
| Class II | 1,016.17 | 1,337.47 | 1,391.42 | 1,166.92 | 1,469.10 |
| Class III | 76.49 | 94.81 | 112.56 | 113.92 | 147.76 |
| Class IV | 0.56 | 2.19 | 3.82 | 5.86 | 6.16 |
| Unclassified | 271.26 | 393.71 | 460.97 | 448.39 | 637.49 |
| Special-class | 4,736.01 | 7,869.72 | 12,949.25 | 13,908.74 | 17,800.90 |

The study recorded an increase in the total expenditure for T2DM treatment in Vietnam, rising from 227.17 million USD in 2018 to 425.53 million USD in 2022. Among the components, drug expenditure consistently accounted for the largest share, ranging from 57.75% to 64.30%. Most categories exhibited an upward trend from 2018 to 2020, experienced a decline in 2021, and then resumed growth in 2022. Additionally, the expenditure for transportation constituted a very small and negligible proportion.

#### Proportion of the Total Expenditure for Healthcare Facilities

**Fig 1.**
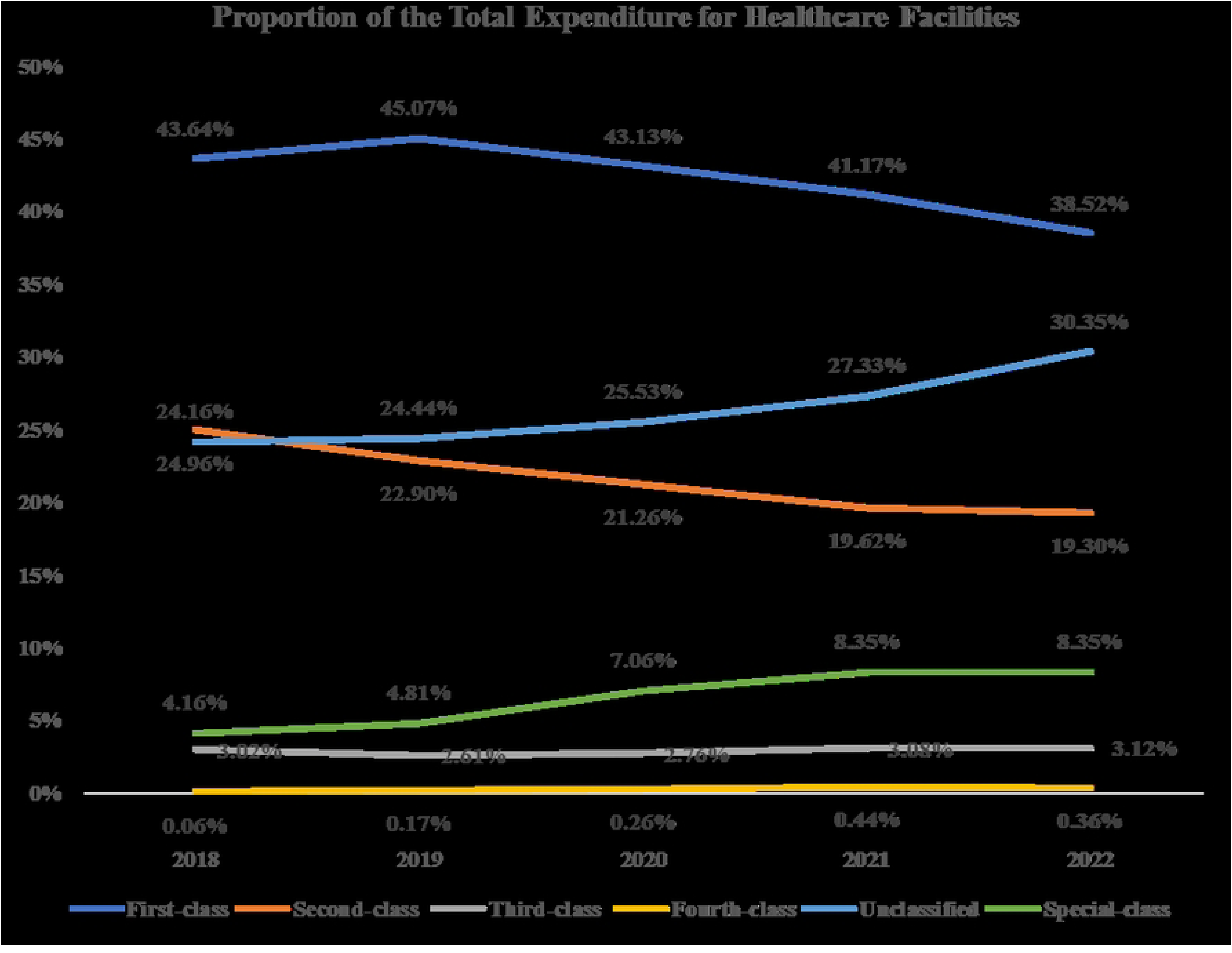
Proportion of the Total Expenditure for Healthcare Facilities.

The study found that Class I healthcare facilities consistently accounted for the largest share of total health expenditure, although this proportion showed a downward trend over the years, declining from 43.64% in 2018 to 38.52% in 2022. Total health expenditure at Class II healthcare facilities represented the second-largest share and also decreased over time, from 24.96% to 19.3%. In contrast, the shares of total health expenditure allocated to unclassified healthcare facilities and special-class facilities increased steadily over the years, from 24.16% to 30.35% and from 4.16% to 8.35%, respectively. Health expenditure at Class III and Class IV facilities accounted for only a negligible proportion, remaining below 5%.

#### Healthcare Facilities class I

The survey of the total expenditure by component in healthcare facilities class I from 2018 to 2022 recorded results, which are presented in **Fig 2**.

**Fig 2.**
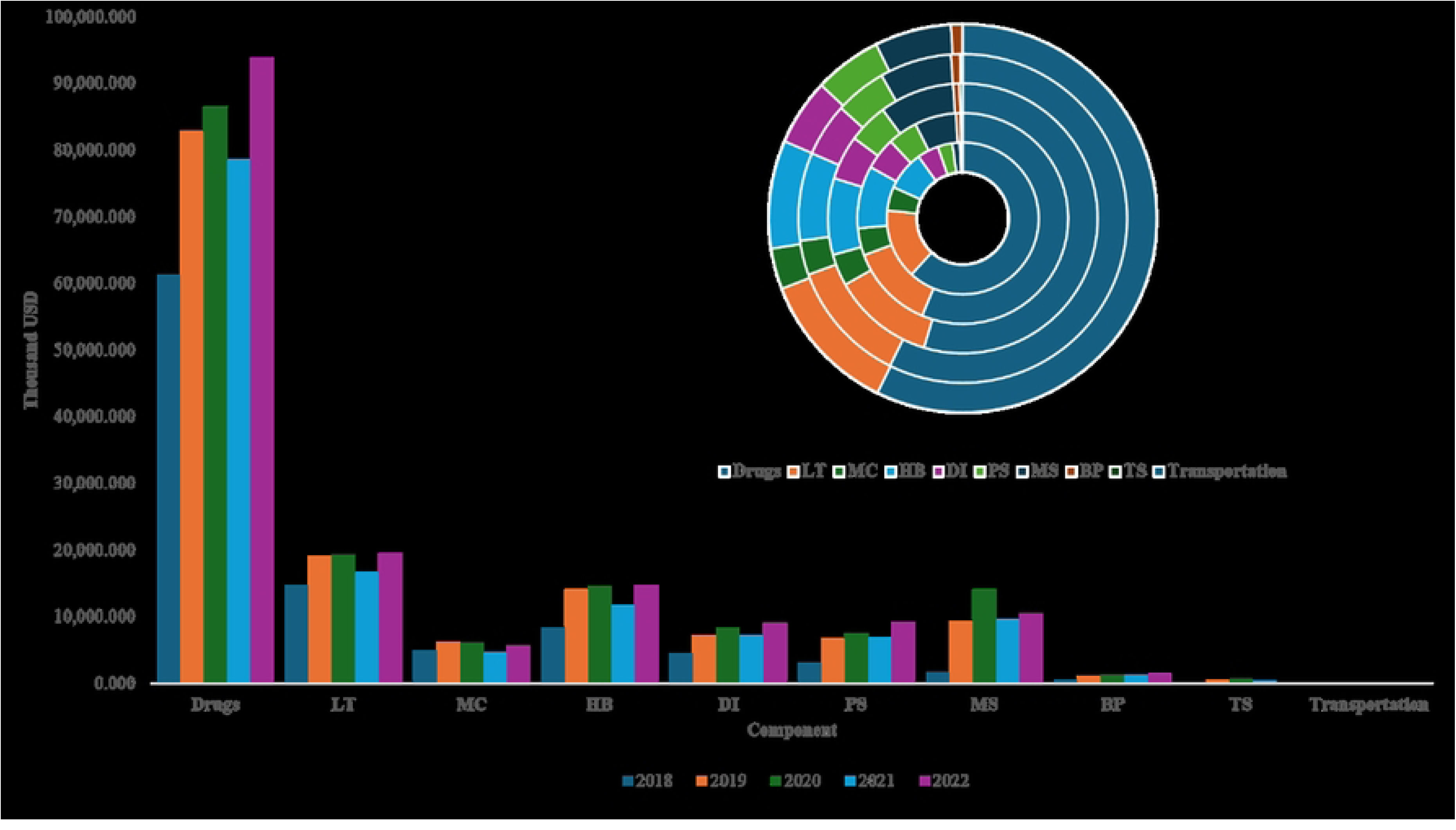
Total Expenditure by Component for Healthcare Facilities class I (Thousand USD) Notes LT: Laboratory Test; MC: Medical Consultation; MB: Hospital bed; DI: Diagnostic Imaging; SP: Procedures and Surgeries; MS: Medical supplies; BP: Blood Products TS: Technical Services.

The results show that the drug component consistently accounted for the highest proportion, ranging from 54.65% to 61.79%, with the expenditure increasing from 61,252.50 thousand USD in 2018 to 78,514.33 in 2022. The laboratory test expenditure ranked second, accounting for 11.86% to 14.90%, increasing from 14,722.73 thousand USD to 16,640.80 thousand USD. The expenditure for medical consultations and hospital beds accounted for 3.40% to 5.05% and 8.38% to 9.59%, respectively, with corresponding increases from 5,008.24 thousand USD and 8,309.20 thousand USD in 2018 to 4,616.80 thousand USD and 11,867.95 thousand USD in 2022. Furthermore, the expenditure for diagnostic imaging ranged from 4.55% to 5.52%, increasing from 4,514.16 thousand USD to 7,186.67 thousand USD. Procedures/surgeries and medical supplies also increased over time, while blood products and technical services remained below 0.93% of total expenditure.

#### Healthcare Facilities class II

The survey of the total expenditure by component in healthcare facilities class II from 2018 to 2022 recorded results, which are presented in **Fig 3**.

**Fig 3.**
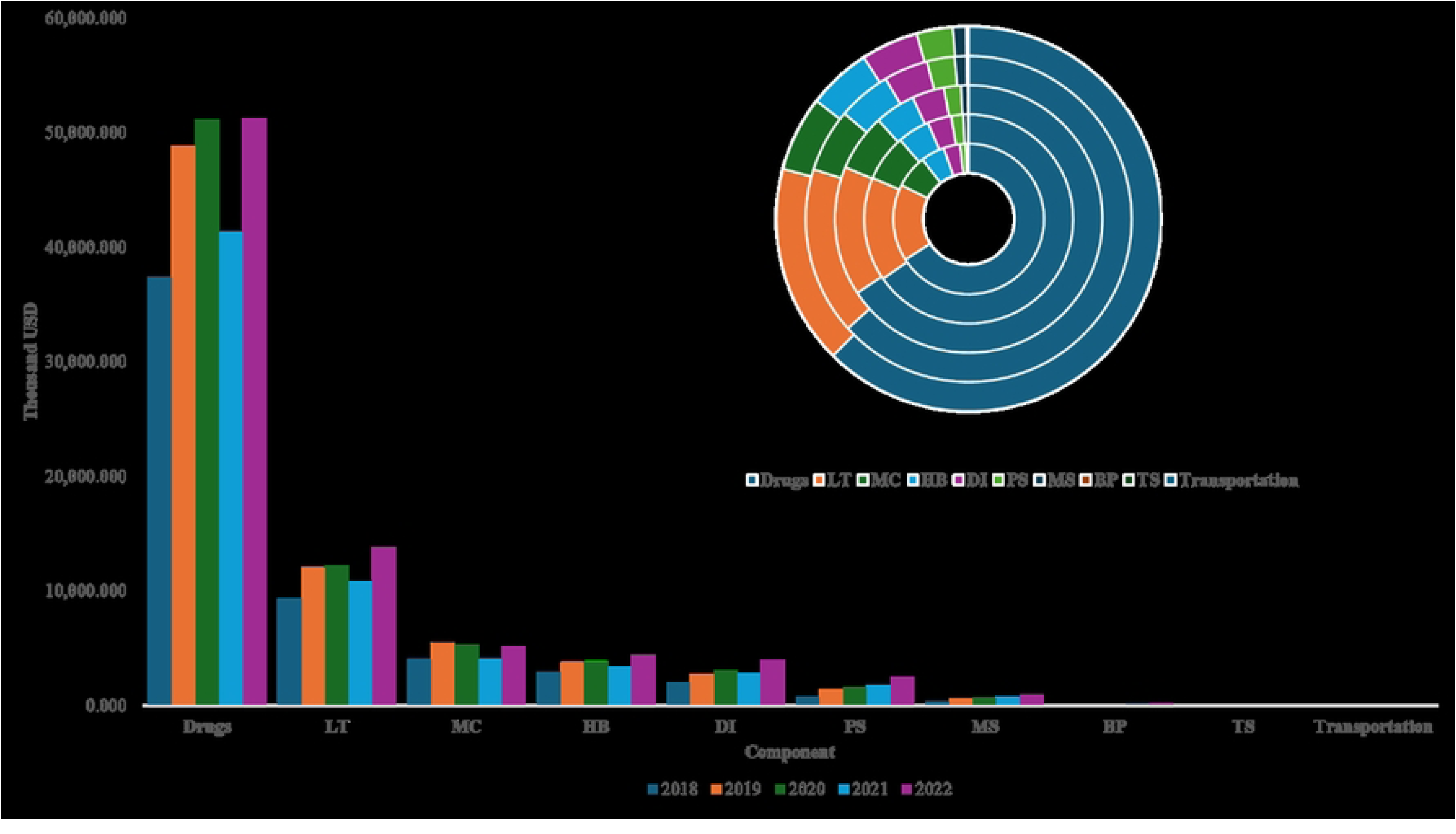
Total Expenditure by Component for Healthcare Facilities class II (Thousand USD) Notes LT: Laboratory Test; MC: Medical Consultation; MB: Hospital bed; DI: Diagnostic Imaging; SP: Procedures and Surgeries; MS: Medical supplies; BP: Blood Products TS: Technical Services.

The survey of the total expenditure by component in healthcare facilities class II from 2018 to 2022 showed that the drug expenditure consistently accounted for the highest proportion, ranging from 62.45% to 65.93%. The drug expenditure increased from 37,387.69 thousand USD in 2018 to 41,340.40 thousand USD in 2022. The laboratory test expenditure ranked second, accounting for 15.64% to 16.76%, increasing from 9,352.50 thousand USD to 13,759.09 thousand USD. The expenditure for medical consultations and hospital beds accounted for 6.26% to 7.21% and 4.93% to 5.33%, respectively, with corresponding increases from 3,998.92 thousand USD and 2,915.25 thousand USD in 2018 to 5,173.38 thousand USD and 4,374 thousand USD in 2022. Additionally, the expenditure for diagnostic imaging accounted for 3.53% to 4.81%, increasing from 2,001.72 thousand USD to 2,867.01 thousand USD. Other components, such as surgical procedures, consumable medical supplies, blood, and transportation, accounted for negligible proportions, each representing less than 3.04% of the total expenditure.

#### Healthcare Facilities class III

The survey of the total expenditure by component in healthcare facilities class III from 2018 to 2022 recorded results, which are presented in **Fig 4**.

**Fig 4.**
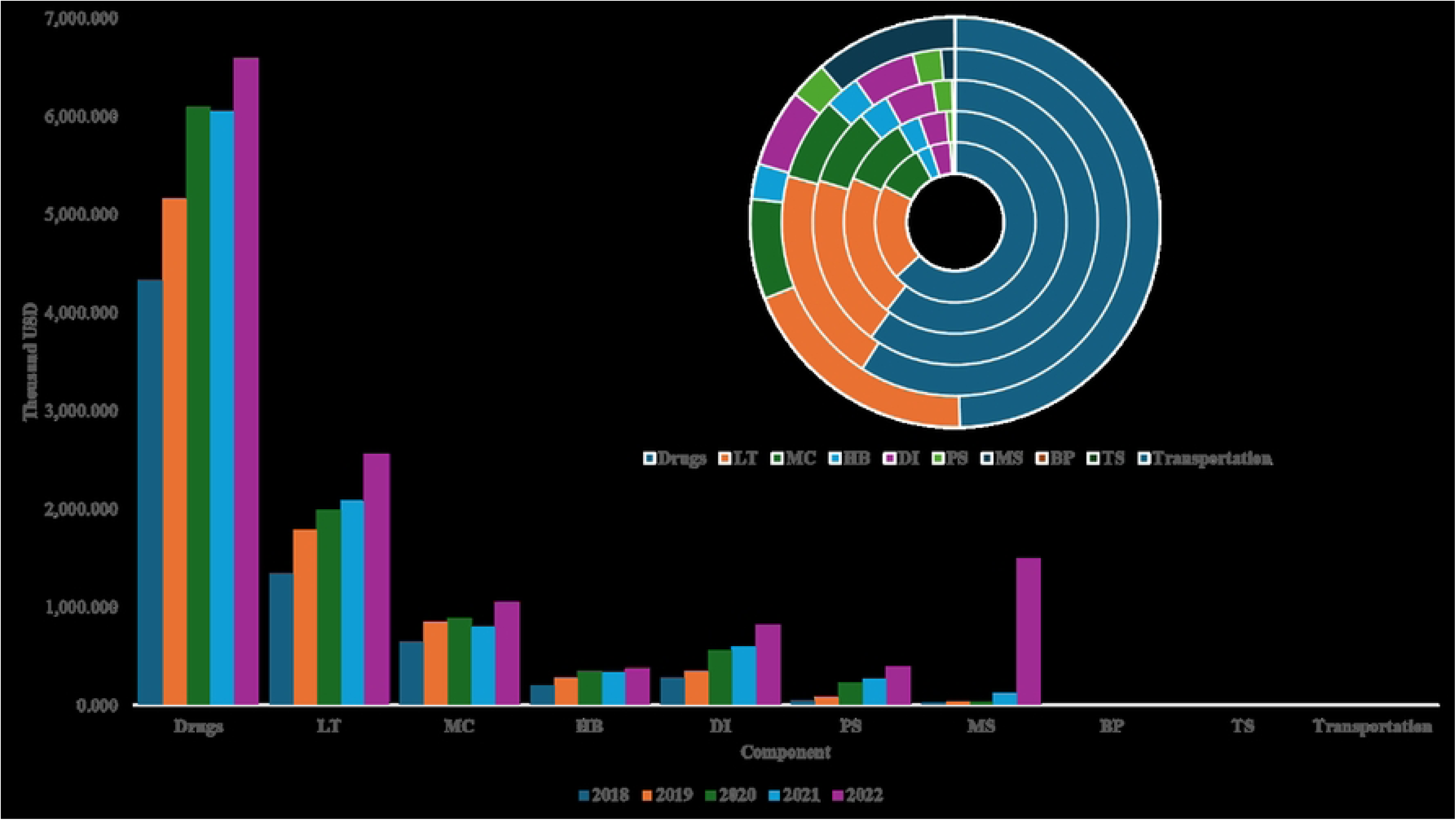
Total Expenditure by Component for Healthcare Facilities class III (Thousand USD) Notes LT: Laboratory Test; MC: Medical Consultation; MB: Medical bed; MID: Medical Imaging Diagnosis; SP: Surgery Procedures; MS: Medical supplies; TS: Technical Services.

The survey of the total expenditure by component in healthcare facilities class III from 2018 to 2022 showed that the drug expenditure consistently accounted for the highest proportion, ranging from 49.65% to 63.10%. The drug expenditure increased from 4,329.05 thousand USD in 2018 to 6,589.67 thousand USD in 2022. The laboratory test expenditure ranked second, accounting for 19.29% to 20.96%, increasing from 1,335.87 thousand USD to 2,560.77 thousand USD. The expenditure for medical consultations and hospital beds accounted for 7.70% to 9.99% and 2.78% to 3.45%, respectively, with corresponding increases from 647.49 thousand USD and 197.89 thousand USD in 2018 to 1,047.64 thousand USD and 369.05 thousand USD in 2022. Additionally, the expenditure for diagnostic imaging accounted for 4.11% to 6.14%, increasing from 281.82 thousand USD to 815.40 thousand USD. Other components, such as surgical procedures, blood, and transportation, accounted for negligible proportions, each representing less than 3% of the total expenditure. Notably, the expenditure for consumable medical supplies showed strong growth throughout the period, increasing from 0.66% to 11.24%, corresponding to an increase from 20.99 thousand USD to 1,492.26 thousand USD.

#### Healthcare Facilities class IV

The survey of the total expenditure by component in healthcare facilities class IV from 2018 to 2022 recorded results, which are presented in Fig 5.

**Fig 5.**
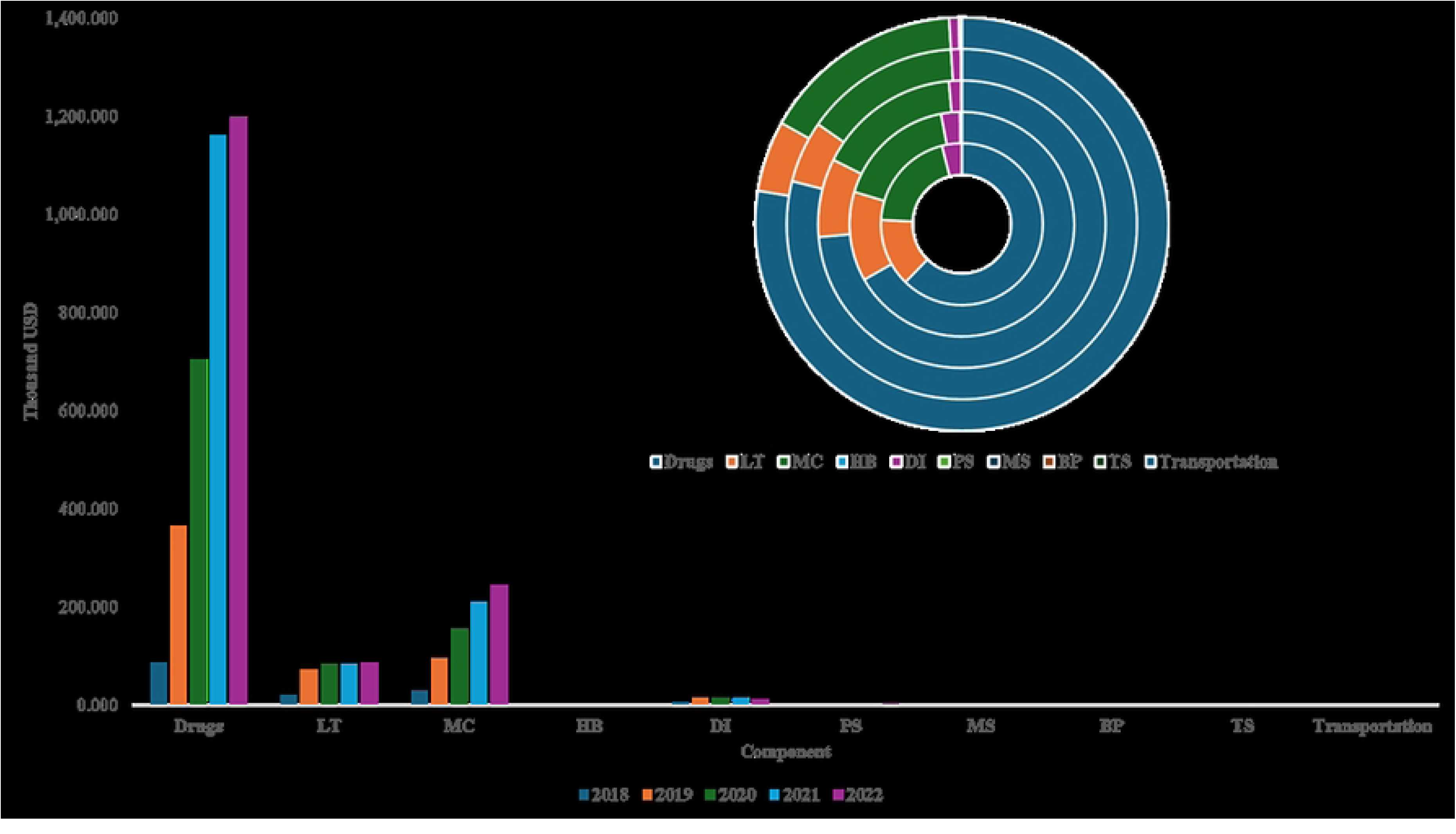
Total Expenditure by Component for Healthcare Facilities class IV (Thousand USD) Notes LT: Laboratory Test; MC: Medical Consultation; MB: Medical bed; MID: Medical Imaging Diagnosis; SP: Surgery Procedures; MS: Medical supplies; TS: Technical Services.

The survey of the total expenditure by component in healthcare facilities class IV from 2018 to 2022 showed that the drug expenditure consistently accounted for the highest proportion, ranging from 62.30% to 78.95%. The drug expenditure increased from 87.78 thousand USD in 2018 to 1,160.54 thousand USD in 2022. The laboratory test expenditure ranked second, accounting for 5.55% to 13.55%, increasing from 19.10 thousand USD to 84.16 thousand USD. Notably, the expenditure for medical consultations increased nearly 10 times, from 28.44 thousand USD in 2018 to 210.73 thousand USD in 2022. Additionally, other components such as medical imaging diagnosis and surgical procedures accounted for negligible proportions, each representing less than 3.94% of the total expenditure.

#### Unclassified Healthcare Facilities

The survey of the total expenditure by component in unclassified healthcare facilities from 2018 to 2022 recorded results, which are presented in **Fig 6**.

**Fig 6.**
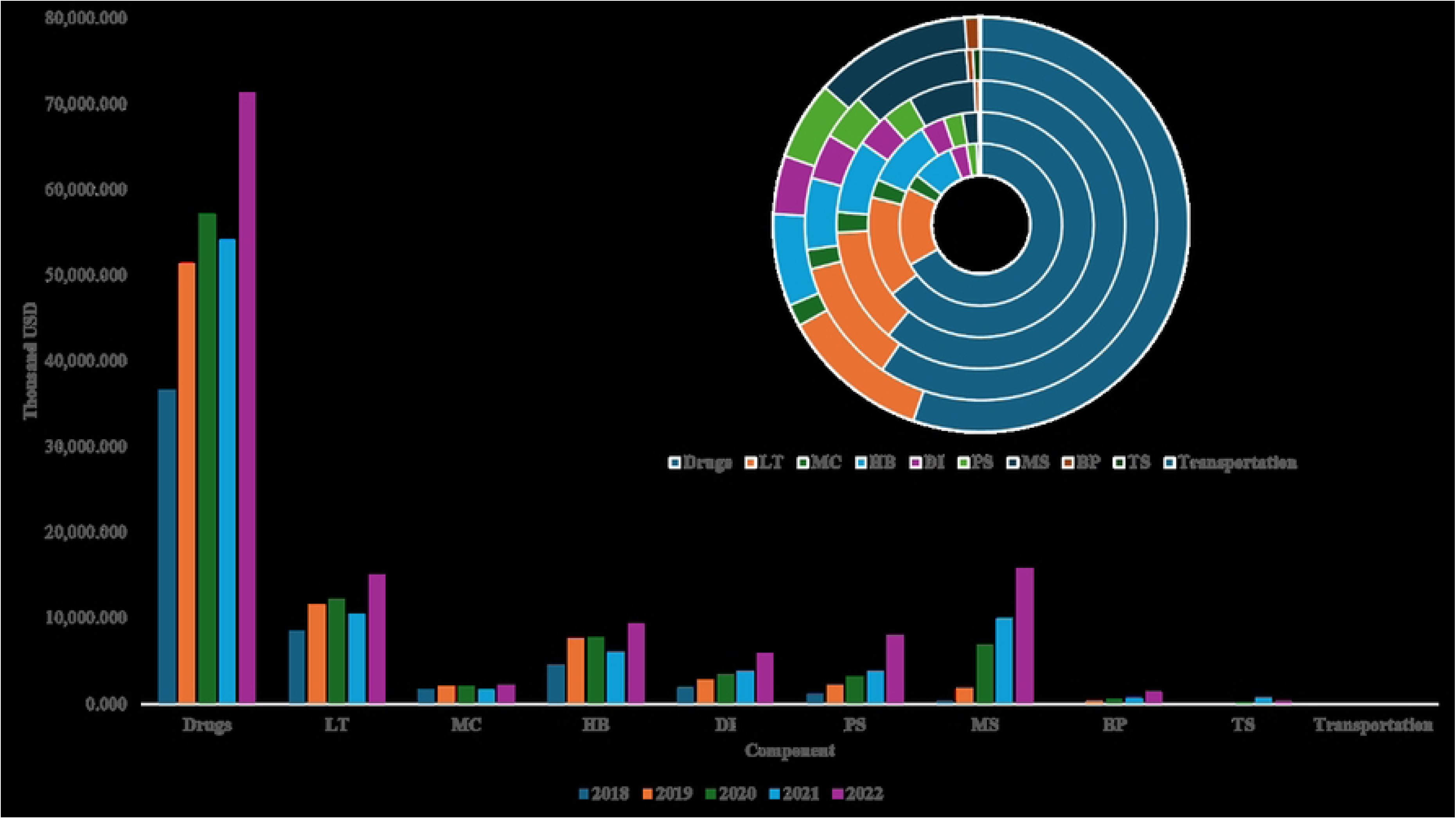
Total Expenditure by Component for Unclassified Healthcare Facilities (Thousand USD) Notes LT: Laboratory Test; MC: Medical Consultation; MB: Medical bed; MID: Medical Imaging Diagnosis; SP: Surgery Procedures; MS: Medical supplies; TS: Technical Services.

The survey on the total expenditure by component at Unclassified healthcare facilities from 2018 to 2022 shows that the pharmaceutical group consistently holds the largest share, ranging from 55.28% to 66.76%, with spending increasing from 36,633.90 thousand USD in 2018 to 71,402.52 thousand USD in 2022. The laboratory testing group ranks second in terms of proportion, ranging from 11.41% to 15.65%, with the expenditure increasing from 8,586 thousand USD to 15,031.65 thousand USD over the same period. The expenditures for outpatient consultations and inpatient bed days account for 1.71% to 3.03% and 7.19% to 9.60%, respectively, with spending increasing from 1,665.36 thousand USD to 2,203.65 for outpatient consultations, and from 4,568.27 thousand USD to 9,292.62 thousand USD for inpatient bed days. Medical imaging accounts for 3.37% to 4.56%, with the expenditure increasing from 1,846.70 thousand USD to 5,894.09 thousand USD. The expenditures for procedures – surgeries and consumable medical supplies show significant growth, increasing from 1,092.75 thousand USD to 7,918.76 thousand USD (share ranging from 1.99% to 6.13%) and from 355,81 thousand USD to 15,808.84 thousand USD (share ranging from 0.65% to 12.24%). Other expenditure groups, such as blood and high-tech medical services, have a small proportion, ranging from 0.15% to 1.08% for blood and from 0.01% to 0.72% for high-tech medical services, with expenditures increasing slightly over the years. Notably, the transport expenditure has hardly incurred any costs during the study period.

#### Special-class Healthcare Facilities

The survey of the total expenditure by component in Special-class healthcare facilities from 2018 to 2022 recorded results, which are presented in **Fig 7**.

**Fig 7.**
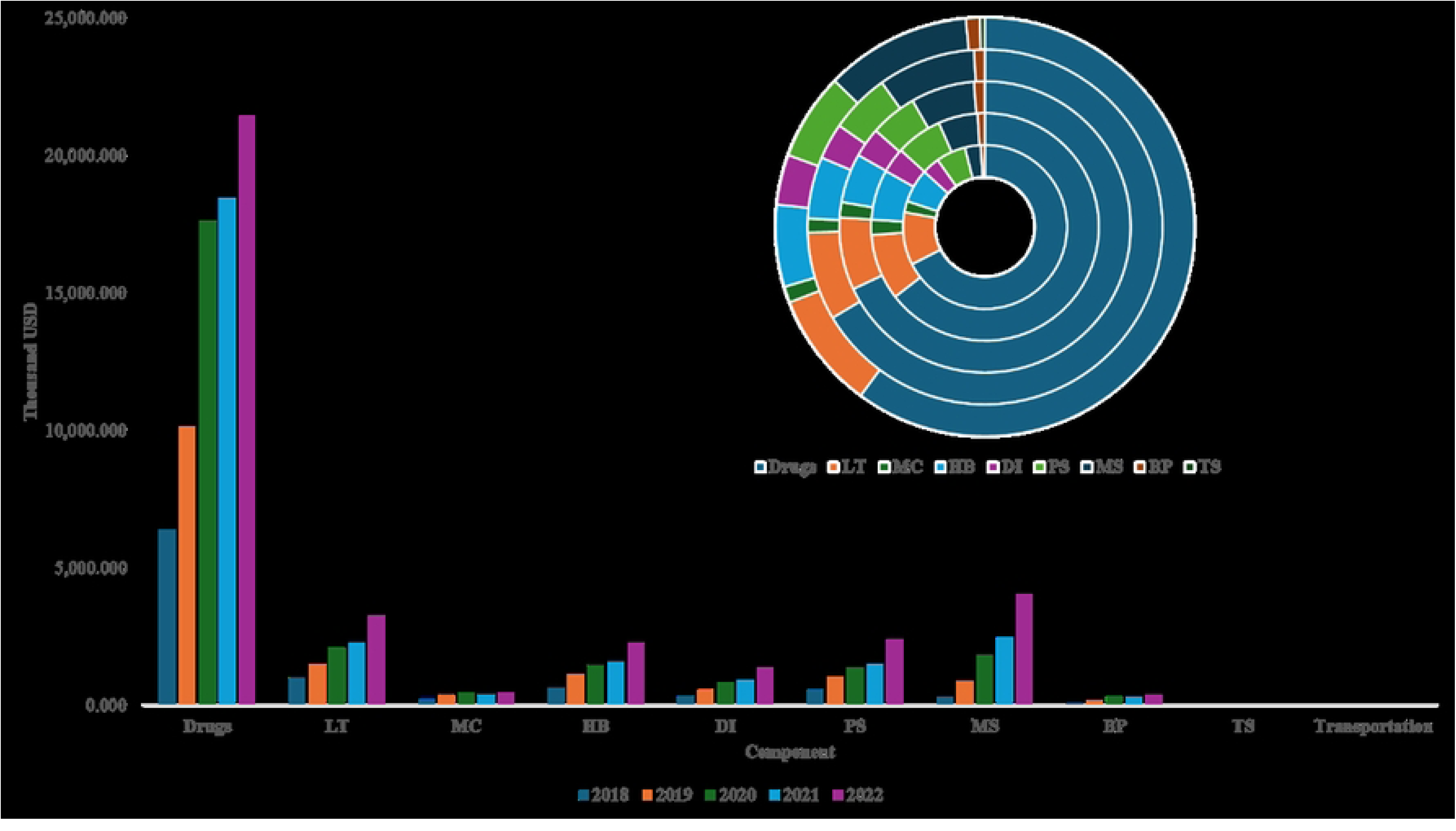
Total Expenditure by Component for Special-class Healthcare Facilities (Thousand USD) Notes LT: Laboratory Test; MC: Medical Consultation; MB: Medical bed; MID: Medical Imaging Diagnosis; SP: Surgery Procedures; MS: Medical supplies; TS: Technical Services.

The survey on the total expenditure by component at special-class healthcare facilities from 2018 to 2022 shows that the drugs group consistently holds the largest share, ranging from 60.34% to 68%, with spending increasing from 6,368.83 thousand USD to 21,441.74 thousand USD in 2022. The laboratory testing group ranks second in terms of proportion, accounting for 8.05% to 10.42%, with the expenditure increasing from 983.84 thousand USD to 3,219.55 thousand USD over the same period.

Additionally, the expenditures for outpatient consultations and inpatient beds accounted for 1.25% to 2.18% and 5.47% to 7.14%, respectively, with spending increasing from 206.11 thousand USD to 445.36 thousand USD for outpatient consultations, and from 629.02 thousand USD to 2,256.68 thousand USD for inpatient beds. The diagnostic imaging expenditure accounted for 3.28% to 3.60%, with the expenditure increasing from 327.02 thousand USD to 1,380.57 thousand USD. The expenditures for procedures - surgeries and medical supplies showed strong growth, increasing from 551.58 thousand USD to 2,392.00 thousand USD (from 5.21% to 6.73%) and from 296.22 thousand USD to 4,017.29 thousand USD (from 3.14% to 11.30%), respectively.

Moreover, the expenditures for other components, such as blood and advanced medical services, represented a small and insignificant proportion, ranging from 0.81% to 1.19% for blood, with the expenditure increasing from 76.87 thousand USD to 381.32 thousand USD, and from 0% to 0.005% for technical services.

#### Average health expenditure for type 2 diabetes treatment at each hospital

Average health expenditure for type 2 diabetes treatment increased over time in most hospital classes from 2018 to 2022. Special-class hospitals had by far the highest expenditure and showed the sharpest rise, from 4,736.01 thousand USD in 2018 to 17,800.90 thousand USD in 2022. First-class and second-class hospitals also recorded relatively high expenditure, although their increases were more moderate. In contrast, class I, class II, and unclassified hospitals accounted for much lower expenditure, despite a gradual upward trend over the study period.

## 4. Discussion

The study showed that the proportion of female patients is higher than that of male patients, and this trend was observed similarly across all of healthcare facilities, except for the special classification, where the ratio was reversed. The study’s findings on the gender distribution of patients across healthcare facility classes were consistent with those reported by Nguyen Thi Thuy Trang et al. (2022), which also found a male-to-female ratio of diabetic patients of 1:1.43 (7). The proportion of female patients showed a gradual decline over the years at class I and unclassified healthcare facilities, and at class II and class III facilities during the period from 2018 to 2021. Similarly, Trung Quang Vo et al. (2018) also reported a decreasing trend in the proportion of female patients from 2013 to 2017 at a private hospital in Ho Chi Minh City (8). This downward trend may reflect improved detection and treatment uptake among men over time, as well as possible changes in health-seeking behavior, referral patterns, or the patient population across healthcare facilities.

Across healthcare facility classes, patients aged 60 years and older accounted for the highest proportion and showed an increasing trend over time. This finding suggests a population ageing trend among patients with type 2 diabetes in the context of rising life expectancy in Vietnam. The results across all healthcare facilities are consistent with those reported in other studies conducted in Vietnam (7–10).

In terms of comorbidities, the primary comorbid conditions of T2DM were DLD and HTN, with the comorbidity rate for both diseases being significantly higher than in other groups. This pattern was also observed across all HFCs tiers. It is evident that T2DM, HTN, and DLD are closely related and have a serious impact on the health of T2DM patients. Additionally, the study analyzed the complications of T2DM, including LBV and SBV complications, metabolic complications, and other complications in the study population from 2018 to 2022. In all tiers of healthcare facilities, the LBV complication group consistently represented a significantly higher proportion compared to the other groups, with the highest rate observed in 2021. This result was consistent with the increase in the group of patients with both HTN and DLD, as well as the proportion of those experiencing complications due to T2DM in 2021. This suggests that during the peak of the pandemic, patients faced difficulties in managing their diabetes and timely screening for complications, which significantly impacted their health.

Drugs accounted for the largest proportion of expenditure across all hospital classifications, consistent with the findings of a study conducted by Thi Thuy Trang Nguyen and colleagues at District 8 Hospital (11). This result indicates that medication expenditure represents the greatest financial burden for patients with type 2 diabetes, as it is a chronic condition requiring lifelong treatment. The expenditure for laboratory tests accounts for the second-largest share after medication, and this expenditure has steadily increased over the years. This finding was previously recorded in a study by Thi Thuy Ha Nguyen and colleagues at Nhan Dan 115 Hospital (12). Regarding the expenditure fluctuations, it can be observed that all component expenditures across HCFs tended to increase from 2018 to 2022. The component expenditures at class I, class II, and Unclassified HCFs experienced a decline in 2021 before recovering and continuing to grow in 2022. In contrast, the expenditures at the remaining HCFs maintained a continuous upward trend throughout the survey period.

The study findings showed that health expenditure for type 2 diabetes treatment remained concentrated mainly in higher-level healthcare facilities, particularly class I facilities, although the proportion of this group declined from 43.64% to 38.52% during the period 2018–2022. This trend may reflect the fact that patients with diabetes continue to mainly utilize services at facilities with higher professional and technical capacity, especially when the disease is prolonged, accompanied by complications, or requires specialized follow-up. A national study in Vietnam by Pham et al. (2020) reported that the direct cost of diabetes was substantial, with approximately 70% of the total cost related to complications (13). This explanation is consistent with the hospital-centric nature of the Vietnamese health system; the World Health Organization has also noted that Vietnam needs to increase investment in primary healthcare to reduce overcrowding at central and tertiary hospitals, while the World Bank has emphasized that the current system remains oriented toward hospital-based care and requires the “right-siting of care” for chronic conditions such as diabetes. A study in Malaysia found that outpatient costs for diabetes treatment were markedly higher at the tertiary level than at the primary level, with provider costs nearly seven times higher and patients’ direct costs about three times higher at tertiary facilities (14).

A major strength of this study is the use of a large administrative dataset from the two largest urban centers in Vietnam, allowing expenditure patterns to be described across multiple hospital classifications rather than within a single institution. Furthermore, the study also provides a component-level breakdown of expenditure, which is useful for identifying where spending is concentrated within insured T2DM care. However, this study has several limitations. First, as a descriptive analysis based on administrative claims data, it did not assess the factors associated with changes in expenditure over time. Second, the analysis was conducted at an aggregated level and did not provide a detailed breakdown of expenditure for individual drugs or specific medical services. Third, although the study described expenditure trends during the study period, it did not include forecasting analyses. Accordingly, the findings are most useful for describing expenditure patterns and may serve as a basis for future studies with more detailed analytic approaches.

## 5. Conclusion

This study provides a descriptive overview of SHI expenditure for T2DM treatment in Hanoi and Ho Chi Minh City from 2018 to 2022. Across hospital classifications, drugs accounted for the largest share of expenditure, while laboratory tests generally represented the second-largest component. In addition, health expenditure remained concentrated in Class I and Class II healthcare facilities, although their shares declined over the study period, while the proportions attributed to unclassified and special-class facilities increased. These findings suggest the need to strengthen diabetes screening, treatment, and follow-up at lower-level healthcare facilities in order to reduce the burden on higher-level hospitals and improve the efficiency of healthcare resource allocation.

## List of abbreviations

DLD: Dyslipidemia.
DMC: Direct medical cost.
HCF: Healthcare Facilities.
HI: Health Insurance.
HTN: Hypertension.
IDF: International Diabetes Federation.
LBV: Large Blood Vessels.
MC: Metabolic Complications.
SBV: Small Blood Vessels.
SHI: Social Health Insurance.
T2DM: Type 2 Diabetes Mellitus.

## Acknowledgments

We would like to express our sincere gratitude to the Ho Chi Minh City and Hanoi Social Health Insurance for their invaluable support and collaboration in providing access to essential data and information, which greatly facilitated the conduct of this research. Their contribution has been crucial to the successful completion of this study.

## Declarations

### Author contributions

TTTN, LVN, NYNV, CDHN and TTHN conceptualized the manuscript. TTTN wrote the original draft. NYNV, CDHN, TTHN, and TTTN supervised the process. NYNV, CDHN, TTHN, and TTTN reviewed and edited the manuscript. All authors read and approved the final manuscript.

### Data availability

The datasets generated by or analyzed during the current study are available from the author, Thuy Thi Thu Nguyen, upon reasonable request.

### Consent for publication

Not applicable

### Human ethics and consent to participate

This study was a secondary analysis of administrative claims/payment data from the Social Health Insurance agencies of Ho Chi Minh City and Hanoi. Access to the dataset was authorized by the respective agencies. The analytic file available to the research team was de-identified before analysis, no direct patient contact occurred, and no intervention was performed. Accordingly, individual informed consent was not applicable for the use of de-identified routine administrative data. For future submission, the manuscript should also report any available institutional ethics approval or formal exemption documentation, if such documentation exists.

